# Quantitative Analysis of Breast Nuclei Morphology for Cancer Diagnosis Using Supervised Machine Learning

**DOI:** 10.1101/2025.08.24.25334307

**Authors:** Zarlish Attique, Sajjid Khan

## Abstract

**Background:** Breast cancer is the most frequently diagnosed malignancy among women worldwide and a major cause of mortality. Early and accurate detection is vital for improving outcomes, yet conventional diagnostic approaches such as mammography, histopathology, and fine-needle aspirate (FNA) cytology can be limited by observer variability and overlapping morphological features. Machine learning (ML) offers a means to improve diagnostic accuracy by capturing subtle patterns in complex datasets.

**Methods:** This study employed the Wisconsin Breast Cancer Diagnostic (WBCD) dataset, comprising 569 FNA cytology cases with 30 quantitative nuclear morphology features. After correlation analysis, 11 predictors were selected to reduce redundancy while retaining diagnostic power. The dataset was split into training and testing sets using an 85:15 stratified approach. Four supervised classifiers were implemented in Python’s scikit-learn library: Random Forest (RF), Multi-Layer Perceptron (MLP), K-Nearest Neighbors (KNN), and Support Vector Classifier (SVC). Models were tuned using GridSearchCV and evaluated using accuracy, precision, recall, and confusion matrices.

**Results:** Exploratory analysis showed malignant tumors exhibited larger nuclear size and higher concavity features than benign tumors. The MLP achieved the best performance (accuracy 0.95, recall 0.91, precision 0.96), misclassifying only two malignant cases. RF and KNN both reached 0.93 accuracy and 0.97 precision but had lower recall (0.85). SVC achieved perfect precision (1.00) but the lowest recall (0.76), misclassifying eight malignant cases.

**Conclusion:** ML models demonstrated reliable classification of breast tumors from cytomorphological features, with the MLP offering the most favorable balance of sensitivity and specificity. These findings highlight the clinical potential of neural network–based models to support early and accurate breast cancer detection.

## I. INTRODUCTION

Breast cancer remains one of the most prevalent cancers worldwide and a leading cause of mortality among women. Early and accurate diagnosis plays a critical role in improving patient survival rates and optimizing treatment strategies. Conventional diagnostic techniques such as mammography, histopathology, and fine-needle aspirate (FNA) cytology have long been used to detect malignancy (1). However, these approaches are often limited by inter-observer variability and the subtlety of morphological differences between benign and malignant lesions (2). Advances in artificial intelligence (AI) and machine learning (ML) have opened new opportunities to enhance diagnostic accuracy by learning complex patterns in medical data that may not be easily discernible to the human eye (3).

The Breast Cancer Wisconsin Diagnostic (WBCD) dataset provides a widely used benchmark for AI-driven breast cancer classification tasks (4). It is derived from digitized images of FNA slides of breast masses, from which thirty morphological features of cell nuclei are computed. These features quantify characteristics such as radius, perimeter, area, texture, compactness, concavity, and symmetry, offering a numerical representation of tumor morphology. The dataset, labeled as benign or malignant based on histopathological confirmation, provides an ideal platform for evaluating machine learning algorithms in cancer diagnosis.

In this study, multiple supervised machine learning classifiers were implemented to predict breast cancer diagnosis from FNA-derived features. Specifically, four models were evaluated: Random Forest Classifier, Multi-Layer Perceptron (MLP), K-Nearest Neighbors (KNN), and Support Vector Classifier (SVC). These models were chosen because they represent a spectrum of approaches including ensemble methods, neural networks, distance-based algorithms, and margin-based classifiers. The models were trained and tested using an 85:15 stratified split of the dataset to preserve class balance, and their performance was assessed through standard evaluation metrics including accuracy, precision, recall, and confusion matrices.

By systematically comparing these algorithms, this work seeks to identify the most effective AI-based approach for breast cancer diagnosis using histology-derived features. Beyond providing insights into the predictive potential of specific classifiers, this research highlights the growing role of machine learning in medical diagnostics, where reliable, reproducible, and accurate tools can support clinicians in early cancer detection and decision-making.

## II. MATERIALS AND METHODS Dataset description

The analysis was performed using the Breast Cancer Wisconsin Diagnostic (WBCD) dataset, which is publicly available at (https://www.kaggle.com/datasets/uciml/breast-cancer-wisconsin-data). The dataset contains 569 patient records generated from digitized fine-needle aspirate (FNA) images of breast tissue. Each record is described by 30 continuous features that capture morphological properties of cell nuclei, including radius, texture, perimeter, area, smoothness, compactness, concavity, concave points, symmetry, and fractal dimension. Each case is labeled as benign (0) or malignant (1), based on histopathological confirmation.

### Data preprocessing

To prepare the data for machine learning models, several preprocessing steps were applied. First, diagnostic labels were mapped to binary format, with malignant tumors assigned as *y = 1* and benign tumors as *y = 0*. Exploratory data analysis was performed to evaluate the distribution of classes and features. Class imbalance was noted, with 357 benign and 212 malignant cases (5). Scatter plots and histograms were generated to study the distribution of morphological characteristics such as *radius_mean, texture_mean*, and *area_mean*. To examine redundancy among features, correlation matrices were computed separately for the mean, standard error (SE), and worst-case groups. Strong positive correlations were observed between *radius_mean, perimeter_mean*, and *area_mean*, suggesting that these features jointly capture tumor size and shape. The correlation with diagnosis was also assessed, where, for instance, *radius_mean* showed a Pearson coefficient of ***r=0*.*73r = 0*.*73*** with malignancy status. Based on these relationships, a reduced feature set was defined to mitigate overfitting while preserving diagnostic power.

### Feature grouping and correlation analysis

Features were grouped into three categories: mean values, standard error (SE), and worst-case values. Pearson correlation analysis was performed within each group, and between features and the diagnosis variable (6). Strong correlations were observed among size-related features such as *radius_mean, perimeter_mean*, and *area_mean*, which also correlated strongly with malignancy. Compactness, concavity, and concave points were also positively associated with malignancy, while *fractal_dimension_mean* showed a weak negative correlation. Based on these results, 11 predictor variables were selected to avoid redundancy and overfitting while retaining discriminative power. The selected features included *radius_mean, perimeter_mean, area_mean, compactness_mean, concavity_mean, concave points_mean, radius_se, area_se, radius_worst, perimeter_worst*, and *area_worst*. Response variable: y ∈ {0,1} where y = 0 → benign, y = 1 → malignant

### Data partitioning

The dataset was split into training and testing subsets using an 85:15 stratified split to preserve class proportions (7). The training set contained 483 records, while the test set contained 86 records. Predictor variables were separated from the outcome variable, creating input (X) and output (y) datasets for both training and testing phases. Let XX denote the feature matrix and yy the diagnostic labels. The split can be represented as: (X, y) → (X_train, y_train), (X_test, y_test), **X**_**_train**_ ∈ □^**(483×11)**,^ **X_test** ∈ □^**^(86×11)**^

### Machine learning models

#### Random Forest Classifier (RFC)

The Random Forest Classifier is an ensemble method that constructs multiple decision trees through bootstrapped sampling and aggregates their predictions via majority voting (8). This approach reduces variance and mitigates overfitting. Each tree split is determined using the Gini impurity index, which is defined as:

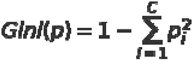

Equation (1): Gini impurity, where p□ = probability of a sample belonging to class I, C = total number of classes

#### Multi-Layer Perceptron (MLP)

The Multi-Layer Perceptron is a feedforward artificial neural network composed of fully connected layers (9). The model was trained using backpropagation and stochastic gradient descent to minimize binary cross-entropy loss:

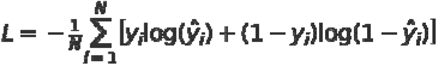

Equation (2): Binary cross-entropy loss, where N = number of training samples y□ = true label of sample i (0 = benign, 1 = malignant) ŷ□ = predicted probability that sample i belongs to class 1

#### K-Nearest Neighbors (KNN)

The KNN algorithm assigns labels to new samples by majority voting among their kk-nearest neighbors in the feature space (10). Similarity between instances is measured using Euclidean distance:

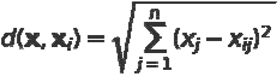

Equation (3): Euclidean distance, where x = query data point, x□ = training data point, n = number of features, x□ = value of feature j for query point, x□□ = value of feature j for training point i

#### Support Vector Classifier (SVC)

The Support Vector Classifier constructs an optimal separating hyperplane by maximizing the margin between classes (11). Given feature vectors xix_i and labels yi∈{−1,1}y_i \in \{**-**1,1\}, the optimization problem is:

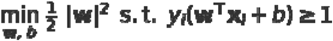

Equation (4): Support Vector Classifier, where w = weight vector (defines the hyperplane), b = bias term (offset of the hyperplane), x□ = feature vector for sample I, y□ = true class label of sample i (+1 or −1)

#### Hyperparameter tuning

Model optimization was performed using **GridSearchCV** with 5-fold cross-validation (12). For the Random Forest model, depth values of 1–4 and tree counts of 10, 50, 100, and 500 were tested. The best parameters identified were a maximum depth of 4 and 50 estimators, which yielded the strongest cross-validation accuracy. Similar experiments were conducted with KNN (varying k values) and MLP (varying hidden layers and iterations) to ensure robust tuning.

#### Model evaluation

Model performance was assessed using accuracy, precision, recall, and confusion matrices. Accuracy measured overall correctness; precision quantified the proportion of predicted malignant tumors that were truly malignant and recall evaluated sensitivity to malignant cases. Confusion matrices were plotted as heatmaps to illustrate true and false predictions for each class. Precision and recall values were also displayed as bar plots for direct comparison across models. These evaluations enabled comprehensive benchmarking of all classifiers under consistent metrics. Overall, this methodology ensured a rigorous preprocessing workflow, careful feature selection, balanced train–test partitioning, systematic model training, and hyperparameter optimization. This framework provided a fair comparison of multiple supervised learning algorithms for diagnosing breast cancer using the WBCD dataset.

## III. RESULTS Exploratory data analysis

Exploratory visualization of the Wisconsin Breast Cancer Diagnostic (WBCD) dataset demonstrated distinct patterns between benign and malignant cases. Scatterplots of nuclear morphology features, such as *radius_mean* versus *texture_mean*, revealed that malignant tumors generally exhibited larger nuclear radii with increased variability. Similarly, the distribution of *area_mean* was markedly right-skewed, with malignant samples concentrated at higher values. Class-colored bivariate plots further emphasized these trends, showing malignant samples clustering toward larger values of *perimeter_se* and *area_se* as well as elevated *smoothness_se* and *compactness_se*. These visualizations confirmed that size and shape descriptors are highly discriminative for malignancy.

### Feature selection and correlation

Correlation analysis highlighted strong collinearity among size-related descriptors (*radius_mean, perimeter_mean, area_mean*) and their “worst” counterparts, consistent with their joint capture of tumor bulk and morphology. Concavity-related features (*concavity_mean, concave points_mean*) also demonstrated strong associations with malignant status. Based on these observations, eleven predictive features were retained for model construction to balance diagnostic power with overfitting risk.

### Model performance

All four machine learning models—Random Forest (RF), Multi-Layer Perceptron (MLP), K-Nearest Neighbors (KNN), and Support Vector Classifier (SVC)—were evaluated on an independent 15% test set. The **Random** Forest achieved an accuracy of 0.93, with high precision (0.97) but moderate recall (0.85). The confusion matrix indicated very few false positives (n = 1), but five false negatives, reflecting occasional missed malignant cases. Examination of individual trees revealed repeated reliance on nuclear size (*perimeter_worst, radius_worst*) and concavity measures for decision splitting. The **MLP** demonstrated the best overall performance, with an accuracy of 0.95 and the highest recall (0.91). Its confusion matrix showed only two malignant cases misclassified as benign, underscoring its sensitivity. The MLP thus offered the strongest trade-off for diagnostic screening, where minimizing false negatives is clinically critical. The **KNN** classifier performed comparably to the RF, with an accuracy of 0.93, precision of 0.97, and recall of 0.85. Like the RF, KNN produced few false positives but missed five malignant cases, suggesting its neighbor-based decision boundaries capture size-related separation but remain less sensitive to subtle malignant morphologies. The **SVC** obtained an accuracy of 0.91 and demonstrated perfect precision (1.00), meaning no benign cases were misclassified as malignant. However, its recall was lowest at 0.76, with eight malignant cases misclassified as benign. This conservative behavior indicates that the linear margin maximized specificity at the expense of sensitivity—a clinically undesirable outcome where missed diagnoses pose significant risks.

### Comparative evaluation

A direct comparison of metrics across models highlighted that the MLP consistently outperformed others in sensitivity, correctly detecting the majority of malignant cases while maintaining high precision. The RF and KNN offered a balanced performance profile, favoring precision over recall, whereas the SVC provided maximal specificity but failed to detect a substantial fraction of malignancies. A direct comparison of classification metrics across models **(Fig. 3)** revealed clear differences in diagnostic behavior. The Multi-Layer Perceptron (MLP) consistently outperformed all other approaches in sensitivity, correctly identifying the majority of malignant cases while maintaining high precision. This balance is particularly critical in a diagnostic setting, where minimizing false negatives—missed cancers—is of greater clinical importance than minimizing false positives. Random Forest (RF) and K-Nearest Neighbors (KNN) displayed comparable profiles, each achieving strong precision (0.97) and overall accuracy (0.93). However, their recall values (0.85) indicated a tendency to misclassify a subset of malignant tumors as benign. This suggests that while both RF and KNN excel at confidently labeling benign cases, their decision boundaries are less sensitive to the morphological heterogeneity observed in malignant nuclei, particularly for cases falling near class margins.

**Fig 1.**
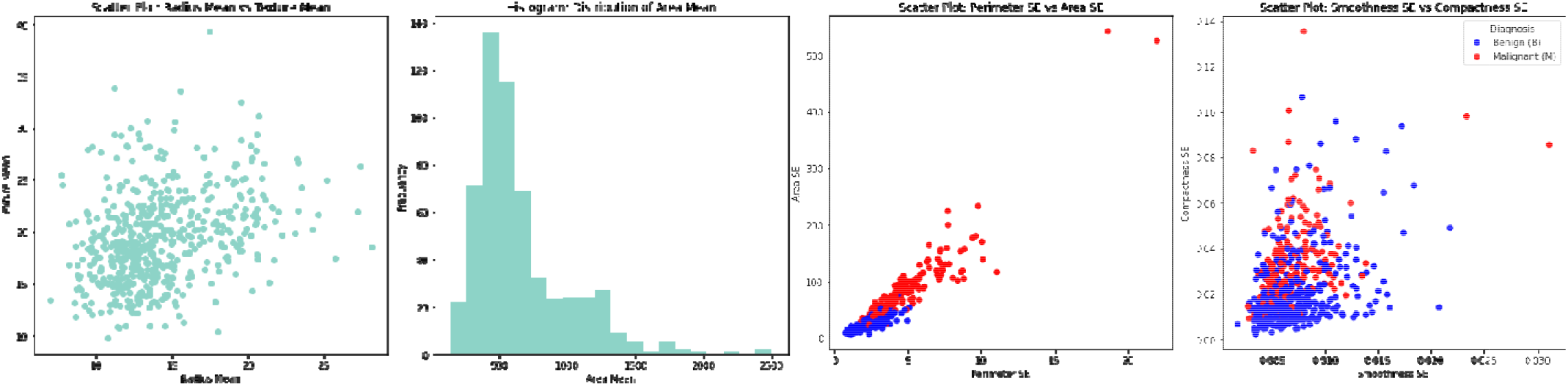
Exploratory plots of nuclear morphology features. Left: scatter of *radius_mean* vs. *texture_mean* demonstrating greater size variation in malignant cases. Right: histogram of *area_mean* showing right-skewed distribution with malignant tumors occupying the upper tail. Class-colored scatter plots illustrating diagnostic separation. Left: *perimeter_se* vs. *area_se*; malignant tumors (red) cluster at higher values than benign (blue). Right: *smoothness_se* vs. *compactness_se* with malignant cases shifted upward/right.

**Fig 2.**
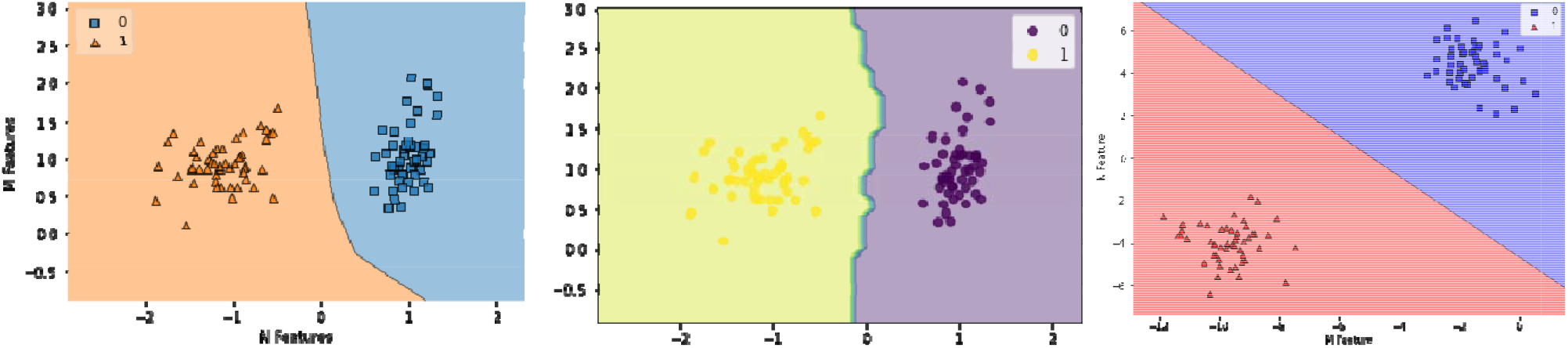
Illustrative decision boundaries for three classifiers trained on synthetic two-feature datasets. (A) Random Forest partitions the feature space with irregular, axis-aligned splits. (B) K-Nearest Neighbors produces piecewise boundaries shaped by local neighbor distributions. (C) Support Vector Classifier establishes a linear separating hyperplane with maximal margin. These examples highlight the distinct boundary geometries that underlie each model’s diagnostic behavior.

**Fig 3.**
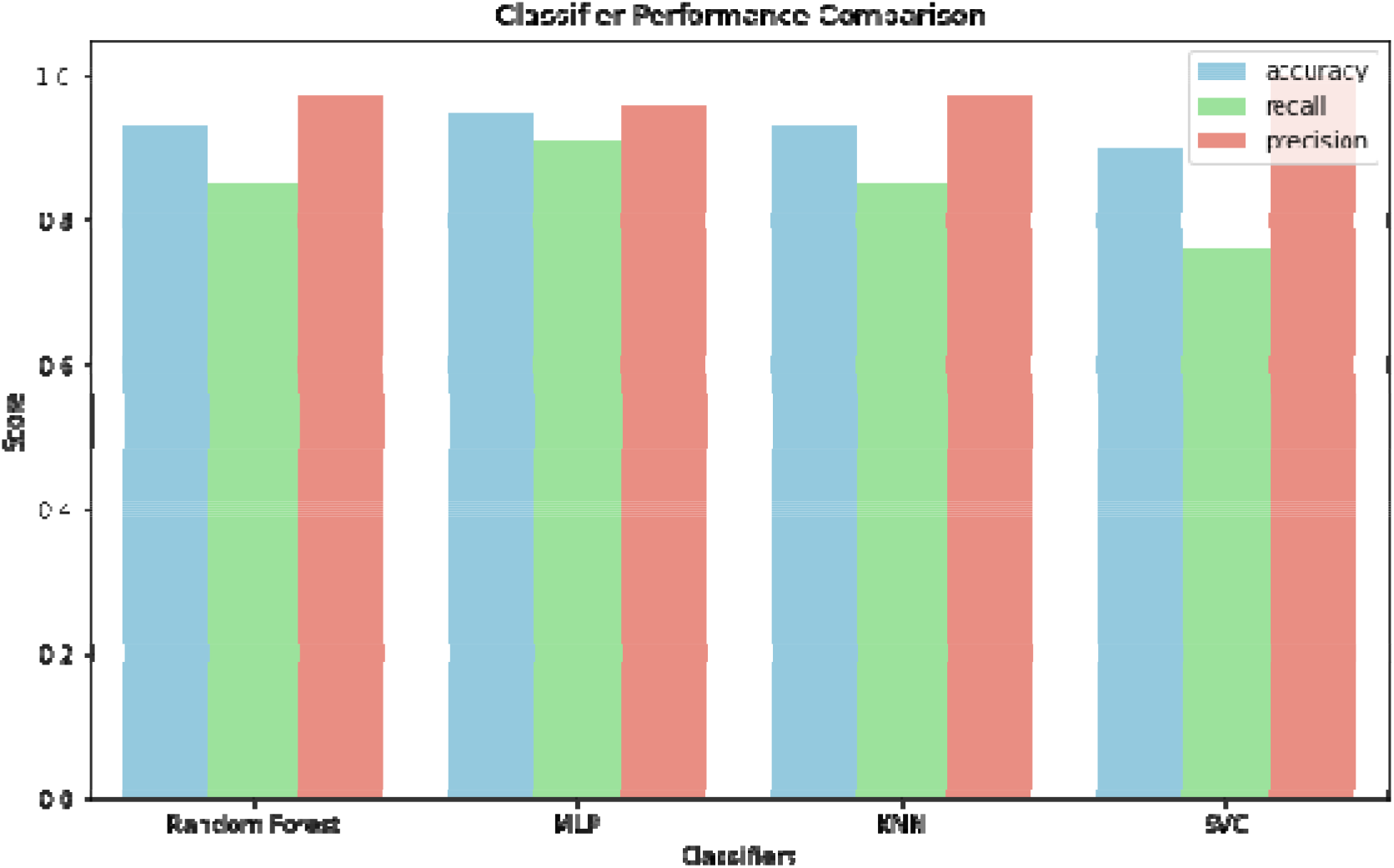
Comparative performance of four classifiers (Random Forest, MLP, KNN, and SVC) evaluated on the WBCD test set. Bars represent accuracy, recall, and precision. The MLP achieved the highest accuracy and recall, the SVC demonstrated perfect precision at the expense of recall, while Random Forest and KNN showed balanced performance with strong precision but moderate sensitivity.

The Support Vector Classifier (SVC) demonstrated the most conservative behavior, achieving perfect precision (1.00) with no false positives, yet at the expense of recall (0.76). This pattern highlights a strict classification margin that prioritizes specificity, resulting in a higher rate of false negatives. While such behavior may be desirable in contexts where false alarms must be minimized, in oncology diagnostics it represents a significant limitation, as undetected malignancies carry substantial clinical risk.

Overall, the comparative evaluation underscores that MLP provides the most clinically relevant balance between sensitivity and precision. RF and KNN represent strong alternatives where interpretability and robustness are prioritized, whereas SVC may be better suited for secondary confirmatory tasks rather than frontline screening. Taken together, these findings reinforce the potential of ML models to supplement diagnostic workflows, while also illustrating the trade-offs inherent in different algorithmic paradigms.

## IV. DISCUSSION

This study demonstrates that nuclear morphological descriptors derived from fine-needle aspirate (FNA) cytology provide highly discriminative information for distinguishing malignant from benign breast lesions (13). Consistent with prior literature, malignant tumors were characterized by increased nuclear size, irregular perimeters, and elevated concavity features, all of which reflect the cytological atypia typical of breast carcinoma. These findings validate the biological relevance of the WBCD dataset and underscore the importance of quantitative nuclear morphology in cancer diagnosis.

The comparative analysis of machine learning models revealed distinct diagnostic behaviors. The Multi-Layer Perceptron (MLP) achieved the highest overall performance, combining excellent accuracy with superior recall (14). Its ability to minimize false negatives is particularly important in clinical oncology, where missing malignant cases poses significant risk to patients. Random Forest and KNN produced comparable accuracy and precision, but their lower recall highlights limitations in capturing the full variability of malignant nuclear features, particularly borderline morphologies. The Support Vector Classifier achieved perfect precision but at the cost of sensitivity, misclassifying several malignant cases. This trade-off suggests that SVC may be more suitable in confirmatory contexts rather than as a primary screening tool.

A key implication of these results is that model selection should be informed not only by statistical performance but also by clinical context. In breast cancer screening and early detection, sensitivity is paramount, making MLP particularly valuable. In contrast, models such as SVC, which prioritize specificity, may have utility in reducing false positives during follow-up diagnostics.

## V. CONCLUSION

Machine learning models provide a reliable framework for classifying breast tumors using cytomorphological features of nuclei. Among the evaluated algorithms, the MLP demonstrated the most favorable balance of accuracy and sensitivity, offering the greatest clinical utility for early detection. RF and KNN remain robust alternatives with high precision, while SVC highlights the trade-off between specificity and sensitivity. Together, these findings reinforce the diagnostic value of nuclear morphological features and support the integration of ML into pathology workflows. Future research should validate these approaches on multi-institutional datasets, incorporate higher-dimensional histological features, and explore deep learning models for end-to-end digital pathology applications.

## DECLARATIONS

### Ethical approval

This study did not involve any experiments on humans or animals conducted by the author. All data analyzed in this study were obtained from publicly available repositories (Kaggle Breast Cancer Wisconsin Diagnostic dataset), which do not require additional ethical approval.

### Consent to participate

Not applicable. The study did not involve human participants, and all data were derived from publicly accessible databases.

### Consent to publish

Not applicable. This manuscript does not contain any individual person’s data in any form (including individual details, images, or videos).

## Acknowledgments

The author would like to thank Government Postgraduate College Mandian, Abbottabad, for their valuable guidance during the course of this work.

## Author’s contributions

Zarlish Attique conceived and designed the study, performed all data analyses and interpretation, acquired and preprocessed the dataset, implemented the machine learning models, visualized the results, and drafted the manuscript.

## Data availability

The Breast Cancer Wisconsin Diagnostic (WBCD) dataset analyzed in this study is publicly available via Kaggle at: https://www.kaggle.com/datasets/uciml/breast-cancer-wisconsin-data. All analysis scripts and processed data are available from the author upon reasonable request.

### Financial support and sponsorship

None.

### Ethical approval and consent to participate

Not applicable.

### Clinical trial number

Not applicable.

